# Predicting 10-year Major Adverse Cardiac Events Using Multi-Source Modalities with XGBoost: Establishing a Baseline for Multimodal Fusion in Cardiac Risk Assessment

**DOI:** 10.1101/2025.08.28.25334683

**Authors:** Roy M. Gabriel, Marly van Assen, Nattakorn Kittisut, Gabrielle Gershon, Xinyue Yan, Carlo N. De Cecco, Ali Adibi

## Abstract

**Background:** Accurate prediction of major adverse cardiac events (MACE) is critical for long-term cardiovascular risk management. Traditional risk scores offer only moderate performance. Leveraging multi-source data may improve individualized risk stratification.

**Methods:** In this retrospective study of patients who underwent non-contrast cardiac-gated CT between 2010 and 2023 across Emory-affiliated hospitals, XGBoost models were trained on structured tabular data using sequential feature integration to predict 10-year MACE. Features included coronary artery calcium (CAC), other imaging-derived metrics, clinical risk scores, electrocardiogram parameters, and laboratory biomarkers. Performance was mainly assessed using AUC-ROC and AUC-PRC. A 5- fold cross-validation strategy was employed, repeated across 10 randomized seeds. Statistical significance was evaluated using two-sided t-tests with 95% confidence.

**Results:** This retrospective study included 25,514 adult patients (mean age 57 ± 10 years; 57% men), of whom 2.93% experienced MACE within 10 years. The final model incorporating all features, achieved the highest performance with an AUC-ROC of 0.883 ± 0.012, a 30.8% improvement over CAC (0.675 ± 0.015), 28.9%-32.2% over clinical risk scores, with p <0.01 for all. AUC-PRC was 0.289 ± 0.028 compared to 0.056-0.104 for clinical risk scores and 0.067 for CAC. SHAP analysis identified creatinine, hemoglobin A1c, body mass index, glomerular filtration rate, and CAC volume as the most influential features.

**Conclusion:** Sequential integration of structured clinical and imaging-derived data significantly improves MACE prediction. This model establishes a robust and interpretable benchmark for future research in multimodal fusion and cardiovascular risk stratification.

## 1. Introduction

Cardiovascular disease (CVD) is the leading cause of death worldwide, accounting for nearly one-third of global mortality^1–3^, with a high prevalence of major adverse cardiovascular events (MACE). Accurate long-term prediction of MACE (e.g., over 10 years) facilitates early intervention, shared decision-making, and allocation of preventative therapies such as lipid-lowering agents, anti-hypertensives, and antiplatelet medications^4^.

The 10-year horizon aligns with established clinical practice guidelines and widely-used risk prediction tools, including the Atherosclerotic Cardiovascular Disease Pooled Cohort Equations (ASCVD PCE risk score)^5–8^ and the Predicting Risk of cardiovascular disease EVENTs (PREVENT) score^9,10^. They leverage demographic, clinical, and laboratory variables to estimate 10-year risk but typically achieve only moderate discriminative performance (AUC-ROC around 0.65-0.75) in large population cohorts. Coronary artery calcium (CAC) scoring on non-contrast cardiac-gated computed tomography (CT) imaging provides direct quantification of subclinical atherosclerosis and has demonstrated incremental value over clinical scores alone^11–15^. Machine learning (ML) approaches have further harnessed tabular clinical data and imaging-derived metrics^16–19^.

Despite these advances, few studies have unified a broad range of relevant features, such as risk scores, demographics, imaging-derived lesion metrics, electrocardiogram (EKG) parameters, and lab biomarkers, into a single framework for MACE prediction in a large, diverse population. Features from different modalities were harmonized into a structured, tabulated format suitable for modeling. Prior work has shown that combining CAC with traditional risk scores improves prediction^17,18^. In this work, we report, for the first time, the integration of demographic variables, clinical risk scores, imaging metrics, EKG parameters, biomarkers, medications, diagnoses, and imaging indications within a ML model, leveraging a large, diverse, and geographically representative dataset to demonstrate superior long-term predictive performance for MACE over existing reports.

In addition to the development of the predictive model, we evaluated whether sequential integration of features can establish a robust baseline for MACE prediction. We trained an extreme gradient boosting (XGBoost)^20^ model on structured multi-source data, quantifying performance gains with each added feature group. We hypothesized that models combining clinical scores, imaging metrics, and biomarkers would outperform single-modality baselines such as ASCVD PCE or CAC score.

## 2. Methods

### 2.1 Dataset

This retrospective study included 25,514 adult patients from Emory Healthcare affiliated hospitals in Atlanta, GA, who underwent non-contrast cardiac-gated CT examination for CAC scoring between 2010 and 2023. Prior history of ASCVD and imaging indication were evaluated based on historical ICD-9 and CPT codes per patient as well as the order report for the CT. This study received ethical oversight from Emory University, and Institutional Review Board approval was granted with a waiver for informed consent due to the retrospective nature.

### 2.2 Features

#### 2.2.1 General

Several modalities were used in this study: demographic data (e.g., age), clinical risk scores (e.g., ASCVD PCE), clinical risk factors (e.g., diabetes), lab results (e.g., lipid panels), medications (e.g., lipid-lowering drugs), EKG parameters (e.g., ventricular rates), and imaging metrics (e.g., CAC volume and density).

#### 2.2.2 CAC Features

CAC score was quantified using the Agatston method, with scores computed by a validated AI algorithm from Siemens Healthineers^21,22^. This AI tool produced accurate and reliable total and vessel-specific CAC scores, closely matching expert consensus. It also reported peak and mean density and lesion counts per vessel (right coronary artery (RCA), left anterior descending artery (LAD), and left circumflex artery).

#### 2.2.3 ASCVD PCE and PREVENT Features

Baseline 10-year ASCVD risk was calculated using both PCE and PREVENT models, originally developed for different age ranges, though all patients were included regardless of age in this study. Additionally, the PREVENT model was used to calculate overall CVD 10- year risk score. Required inputs included blood pressure, cholesterol levels, glomerular filtration rate (GFR), body mass index (BMI), and diabetes status (determined via ICD-9 codes and medication), while optional PREVENT variables, like hemoglobin A1c (HbA1c), were not used.

#### 2.2.4 Clinical Risk Features

The assessed risk factors included hypertension, hypercholesterolemia, hypertriglyceridemia, diabetes, family history of heart disease, and smoking. These were identified using a combination of laboratory results, medication records, and historical ICD-9 diagnostic codes. Hypercholesterolemia, for instance, was defined as having low-density lipoprotein cholesterol ≥120 mg/dL or lipid-lowering medication.

Additionally, prior ASCVD history and the indication for performing cardiac CT were determined using ICD-9/CPT codes and information from the original CT order reports.

#### 2.2.5 EKG Features

Only EKG data within one year of the exam date was included. Extracted features included ventricular and atrial rates, QRS and QT intervals, P, QRS, and T axes, PR interval, waveform onset and offset timings, QRS count, and test reason (e.g., syncope).

#### 2.2.6 Events

Clinical events were determined based on patient-specific ICD-9/10 and CPT codes. MACE was defined as the occurrence of stroke, myocardial infarction, percutaneous coronary intervention, coronary artery bypass grafting beyond 90 days post-CT, or all-cause mortality. MACE served as a critical endpoint in both clinical trials and risk stratification studies. To ensure accuracy, diagnostic code-based event classifications were validated through manual review of electronic health records in over 10% of the cohort, including at least 10% representation from each event category and the non-event group.

### 2.3 Model

An XGBoost model was trained on various sets of medical features, starting with baseline inputs including CAC alone, the ASCVD PCE risk score, and the total ASCVD and CVD PREVENT risk scores. For simplicity, we will subsequently refer to these risk scores as PCE and PREVENT, respectively. Four enhanced feature sets were then developed: IMAGE (other imaging metrics like LAD Agatston score), CR (clinical risk factors, demographics, and physiological variables without imaging), CR+EKG (EKG features), and ALL (all previous features with lab results, medications, diagnoses, and clinical indications). Missing data was not imputed to avoid bias, categorical variables were one-hot encoded, and numerical features were standardized or normalized based on their distribution.

XGBoost was trained with a log loss function and class-weighted loss to address class imbalance. The data was split into training/validation (80%) and testing sets (20%), maintaining consistent MACE distribution across these subsets and ensuring no patient overlap between them. XGBoost was chosen for its strong predictive performance, ability to handle diverse data types, interpretability, and computational efficiency.

We used 5-fold cross-validation on 80% of the data, reserving 20% for final testing. To optimize performance, a grid search over various hyperparameter combinations was performed. The combination yielding the highest average F1-score was selected. This entire process was repeated 10 times with different random seeds to ensure robustness.

### 2.4 Evaluation Metrics

Model performance was evaluated using multiple metrics, including AUC-ROC, AUC-PRC, F1-score, sensitivity, specificity, precision, brier score, and concordance index (C-index). Metrics were summarized across random seeds using the mean and standard deviation. Statistical significance was assessed using two-sided one-sample t-tests against appropriate null hypotheses. All analyses were conducted in Python 3.10.

### 2.5 SHAP Feature Importance Analysis

To assess feature contributions and improve interpretability, SHAP (SHapley Additive exPlanations)^23^ was used on the final XGBoost models. SHAP values quantify the marginal impact of each feature on individual predictions. The mean absolute SHAP values were computed across 10 random seeds to rank features by overall importance and visualize their influence on MACE prediction.

### 2.6 Calibration Curve Analysis

Model calibration was evaluated using reliability plots across 15 probability bins comparing predicted MACE risk with observed outcomes. Given the imbalanced prevalence, the reference was the actual event proportion, ensuring higher predicted risk corresponded to higher observed prevalence.

## 3. Results

### 3.1 Data

The cohort included 25,514 adult patients with a mean age of 56.7 ± 10.1 years, of whom 57.7% were male. Among patients who experienced MACE (n = 747, 2.93%), the mean age was 62.6 ± 11.0 years, with 62.0% being male. The cohort was predominantly White (74.8%), followed by Black (10.6%) and Asian (4.2%) individuals. Additional demographic and clinical characteristics are summarized in Table 1.

**Table 1:** Baseline characteristics for all patients. Continuous variables are reported as mean ± standard deviation. Categorical variables are reported as number and percentage. Comparisons between MACE and No-MACE groups were performed using appropriate statistical tests.

|  | All Patients | MACE Patients | No-MACE Patients | p-value |
| --- | --- | --- | --- | --- |
| **Age** | 56.72 ± 10.06 | 62.60 ± 11.00 | 56.55 ± 9.98 | <0.001 |
| **Gender: Male** | 16,157 (57.7%) | 485 (62.0%) | 15,672 (57.6%) | 0.045 |
| **Race: White** | 20,937 (74.8%) | 595 (76.1%) | 20,342 (74.8%) | 0.437 |
| **Race: Black** | 2,963 (10.6%) | 142 (18.2%) | 2,821 (10.4%) | <0.001 |
| **Race Asian** | 1,169 (4.2%) | 27 (3.5%) | 1,142 (4.2%) | 0.348 |
| **BMI** | 27.99 ± 5.51 | 28.56 ± 6.35 | 27.97 ± 5.48 | 0.011 |
| **Creatinine** | 1.10 ± 1.07 | 1.86 ± 2.70 | 1.06 ± 0.91 | <0.001 |
| **HbA1c** | 8.03 ± 14.88 | 5.92 ± 1.08 | 8.16 ± 15.30 | <0.001 |
| **Troponin-I** | 0.32 ± 2.34 | 0.79 ± 4.55 | 0.19 ± 1.01 | 0.026 |
| **GFR > 70** | 5,948 (35.7%) | 302 (40.5%) | 5,646 (35.4%) | <0.01 |
| **Lipoprotein A** | 39.41 ± 44.19 | 47.38 ± 42.92 | 39.09 ± 44.23 | 0.200 |
| **CAC** | 112.17 ± 339.75 | 416.85 ± 744.15 | 104.13 ± 318.38 | <0.001 |
| **Risk Hypertension** | 13,137 (46.7%) | 609 (77.9%) | 12,528 (45.8%) | <0.001 |
| **Risk Diabetes** | 3,421 (12.2%) | 227 (29.0%) | 3,194 (11.7%) | <0.001 |

### 3.2 Performance Overview

Fig. 1 and Fig. 2 summarize the ROC and PRC curves across 10 random initializations for each feature set. The ALL model demonstrated the strongest discriminative performance, achieving a mean AUC-ROC of 0.883 ± 0.012 and AUC-PRC of 0.289 ± 0.028, outperforming PCE (AUC-ROC = 0.685 ± 0.020), PREVENT (0.668 ± 0.054), and CAC (0.675 ± 0.015) baselines (p < 0.01 for all). CR and CR+EKG yielded moderate gains, with AUC-ROC values of 0.760 ± 0.017 and 0.753 ± 0.016, respectively. The IMAGE model performed poorest (AUC-ROC = 0.650 ± 0.032), with high variability across seeds.

**Fig. 1:**
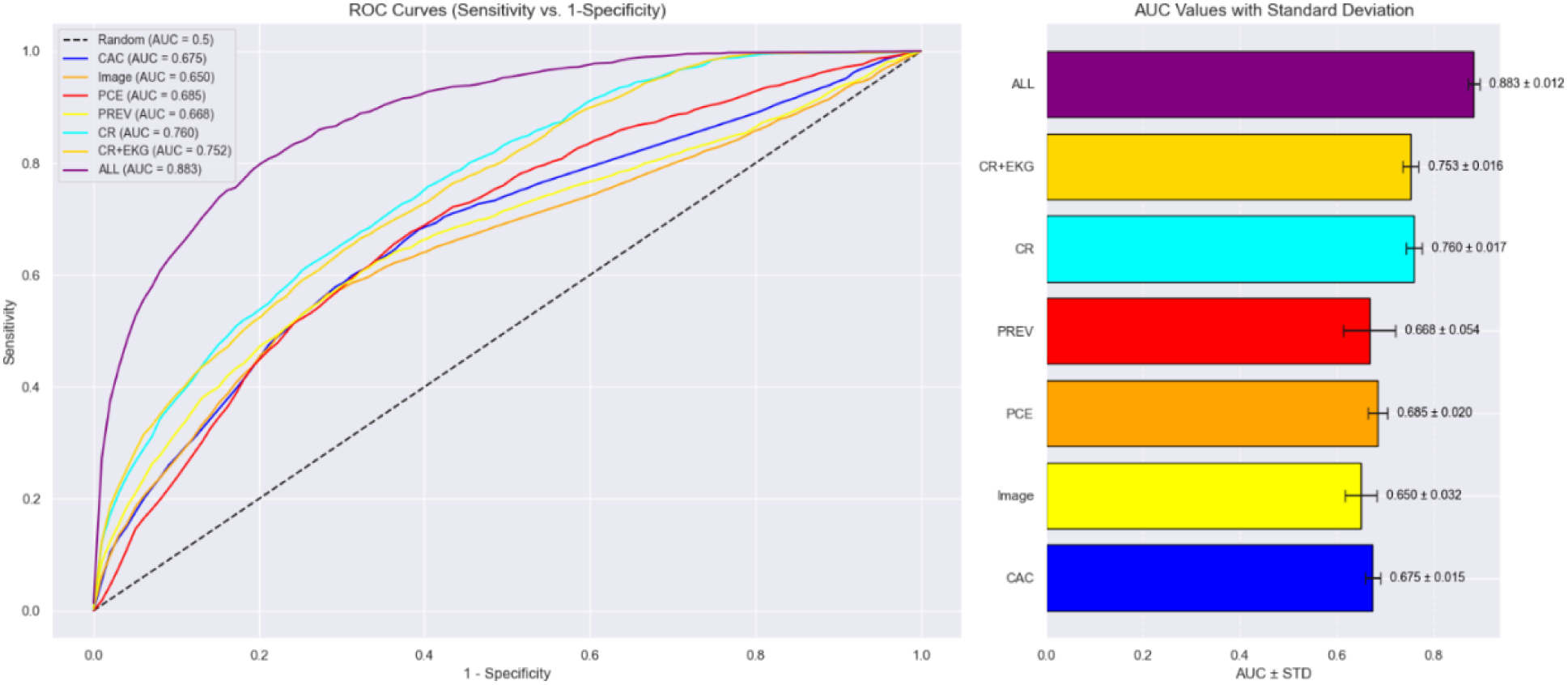
Average AUC-ROC curves across different feature sets (left) and their mean and standard deviation (right).

**Fig. 2:**
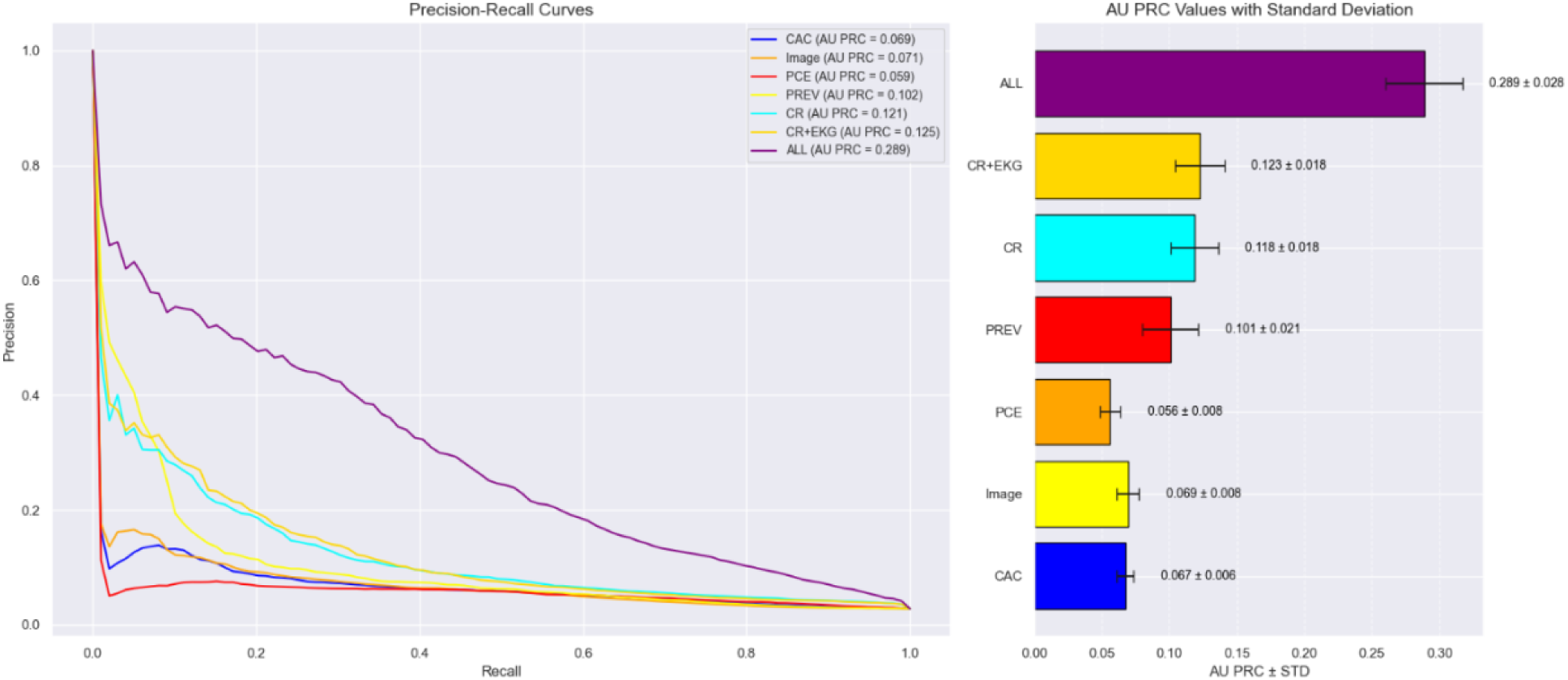
Average AUC-PRC curves across different feature sets (left) and their mean and standard deviation (right).

Table 2 summarizes additional performance metrics across feature sets. The ALL model consistently outperformed others, achieving the highest precision (0.41 ± 0.03), F1- score (0.35 ± 0.03), C-index (0.88 ± 0.01), and the lowest Brier score (0.03 ± 0.00), while maintaining high specificity (0.99 ± 0.00). In contrast, CAC and IMAGE demonstrated lower precision (0.05-0.06), moderate sensitivity (0.47-0.57), and higher Brier scores (0.24 ± 0.02 and 0.21 ± 0.03, respectively). The CR and CR+EKG models showed gains in precision and C-index, but only the ALL model demonstrated consistently strong performance across all metrics, aside from a sensitivity trade-off. All AUC-ROC results were statistically significant (p < 0.01).

**Table 2:** Performance metrics for sequential feature addition. Results are reported as mean (± standard deviation).

|  | ACCURACY | SENSITIVITY | SPECIFICITY | PRECISION | C-INDEX | F1 | BRIER |
| --- | --- | --- | --- | --- | --- | --- | --- |
| **CAC** | 0.71 ($\pm 0.01$) | 0.57 ($\pm 0.03$) | 0.72 ($\pm 0.01$) | 0.05 ($\pm 0.00$) | 0.67 ($\pm 0.02$) | 0.10 ($\pm 0.00$) | 0.24 ($\pm 0.02$) |
| **IMAGE** | 0.78 ($\pm 0.02$) | 0.47 ($\pm 0.06$) | 0.78 ($\pm 0.03$) | 0.06 ($\pm 0.00$) | 0.65 ($\pm 0.03$) | 0.10 ($\pm 0.00$) | 0.21 ($\pm 0.03$) |
| **PCE** | 0.68 ($\pm 0.02$) | **0.60 ($\pm 0.03$)** | 0.68 ($\pm 0.02$) | 0.05 ($\pm 0.00$) | 0.69 ($\pm 0.02$) | 0.09 ($\pm 0.01$) | 0.23 ($\pm 0.01$) |
| **PREVENT** | 0.79 ($\pm 0.03$) | 0.48 ($\pm 0.10$) | 0.80 ($\pm 0.04$) | 0.06 ($\pm 0.00$) | 0.67 ($\pm 0.05$) | 0.11 ($\pm 0.01$) | 0.22 ($\pm 0.02$) |
| **CR** | 0.96 ($\pm 0.00$) | 0.17 ($\pm 0.03$) | 0.98 ($\pm 0.00$) | 0.20 ($\pm 0.04$) | 0.76 ($\pm 0.02$) | 0.18 ($\pm 0.03$) | 0.20 ($\pm 0.04$) |
| **CR+EKG** | 0.96 ($\pm 0.00$) | 0.19 ($\pm 0.02$) | 0.98 ($\pm 0.00$) | 0.20 ($\pm 0.03$) | 0.75 ($\pm 0.02$) | 0.19 ($\pm 0.03$) | 0.20 ($\pm 0.04$) |
| **ALL** | **0.97 ($\pm 0.00$)** | 0.31 ($\pm 0.04$) | **0.99 ($\pm 0.00$)** | **0.41 ($\pm 0.03$)** | **0.88 ($\pm 0.01$)** | **0.35 ($\pm 0.03$)** | **0.03 ($\pm 0.00$)** |

### 3.3 SHAP Feature Importance

Fig. 3 presents SHAP values for the ALL feature model. Features are ranked by their mean absolute SHAP values, with the top five contributors being creatinine, HbA1c, BMI, GFR, and CAC volume (Table 3). Higher values of creatinine, HbA1c, and BMI were associated with increased predicted MACE risk. CAC volume, calcium, and Troponin-I also demonstrated positive SHAP impact. The distributions across other features such as PCE and PREVENT scores were narrower. Interestingly, adding EKG features had minimal impact on feature importance ranking.

**Fig. 3:**
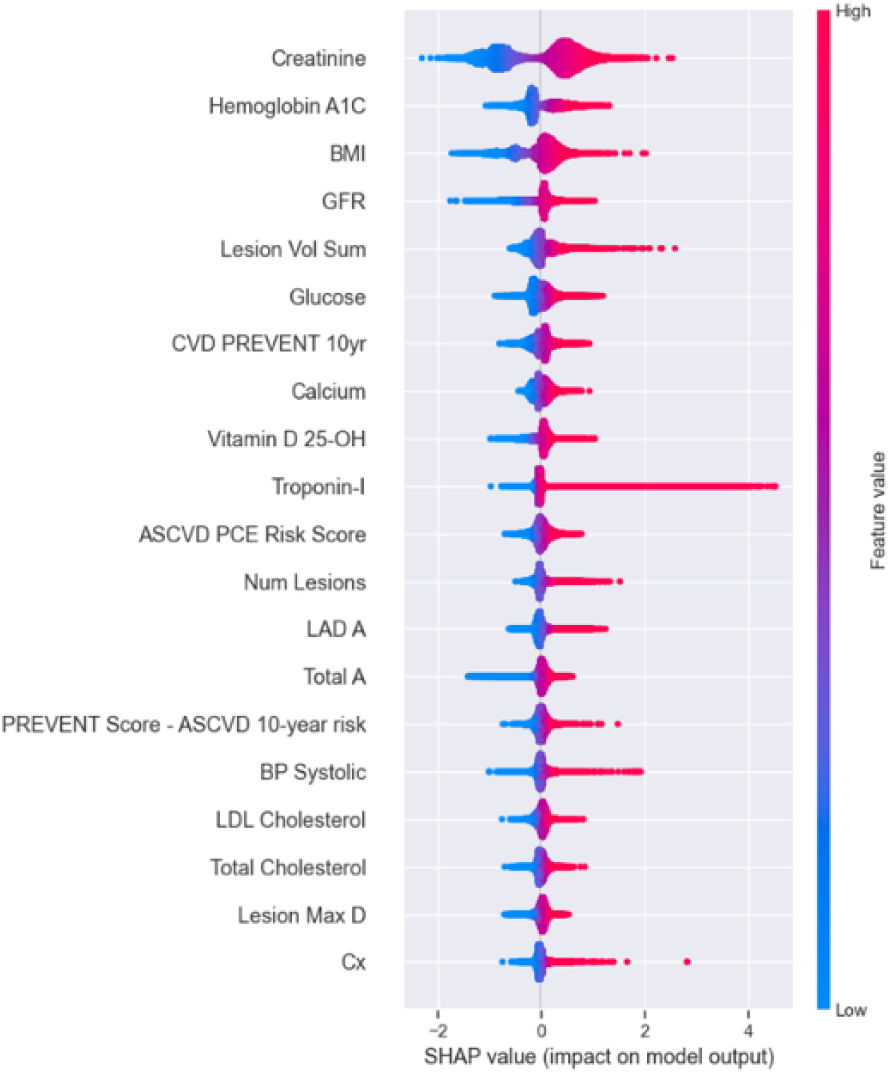
SHAP values across 10 experiments for the ALL feature model.

**Table 3:** Top 5 features for each feature set based on mean absolute SHAP values.

| TOP FEATURES (SORTED BY MEAN ABSOLUTE SHAP) |  |  |  |  |  |
| --- | --- | --- | --- | --- | --- |
| **CAC** | CAC score |  |  |  |  |
| **IMAGE** | CAC Volume | CAC score | Lesion Count | Lesion Max Density | RCA CAC score |
| **PCE** | ASCVD PCE |  |  |  |  |
| **PREVENT** | CVD PREVENT | ASCVD PREVENT |  |  |  |
| **CR** | BMI | CVD PREVENT | ASCVD PCE | ASCVD PREVENT | Hypertension Risk |
| **CR+EKG** | BMI | CVD PREVENT | ASCVD PCE | ASCVD PREVENT | Age |
| **ALL** | Creatinine | HbA1c | BMI | GFR | CAC volume |

### 3.4 Calibration Curves

As shown in Fig. 4, the ALL feature model demonstrated the most accurate calibration among all feature sets, with predicted probabilities closely matching observed MACE rates. It showed a consistent increase in true event frequency across probability bins and aligned most closely with the ideal diagonal reference line. While minor underestimation was observed in lower-risk groups, the model’s predictions became increasingly well-calibrated at higher probability thresholds. In contrast, feature-limited models such as CAC, IMAGE, and PCE exhibited greater deviation from perfect calibration, systematically underestimating risk across most bins.

**Fig. 4:**
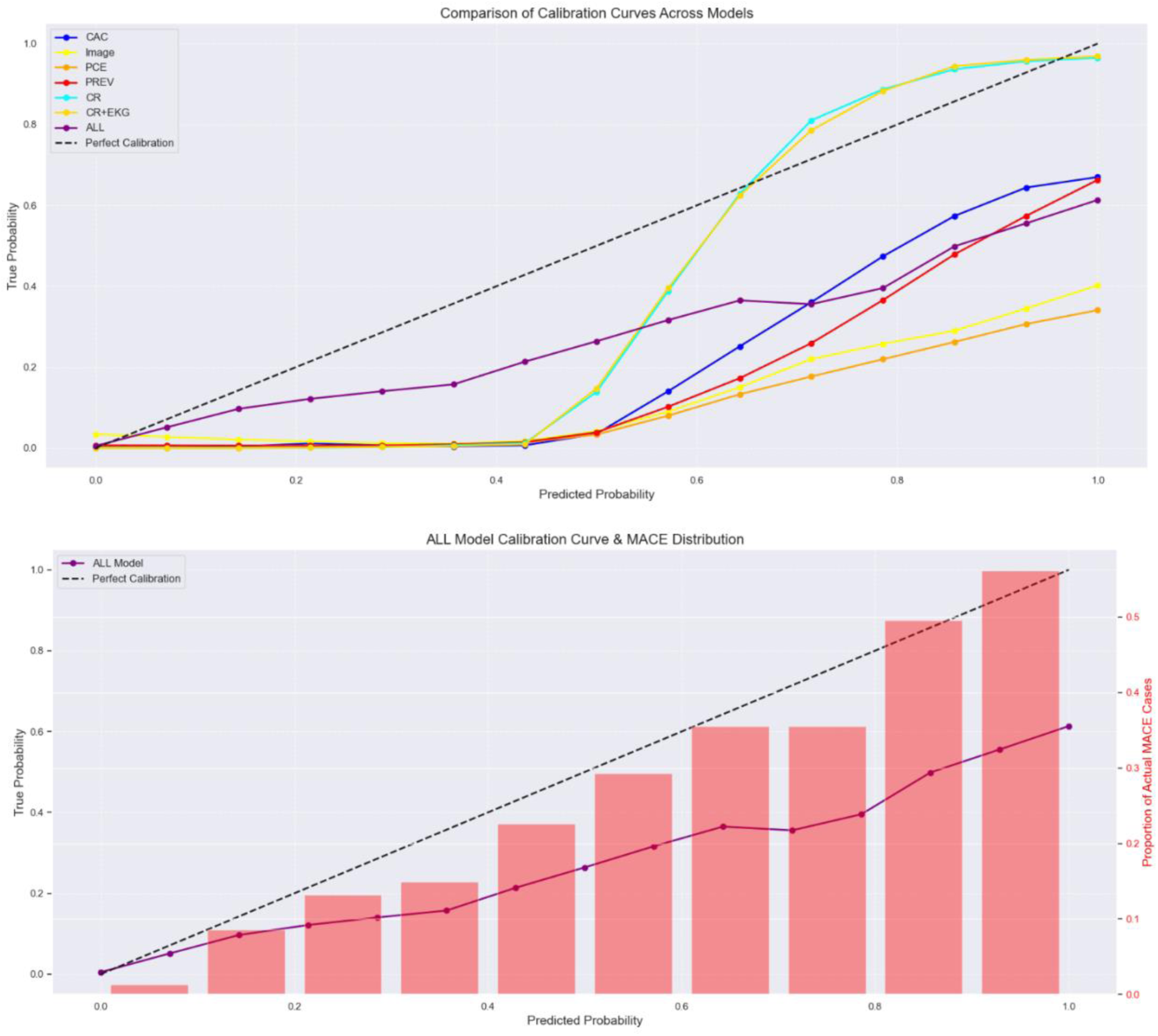
Calibration curves of models trained with different feature sets (up) showed that the ALL feature set exhibited the best overall calibration. Further, the ALL feature model with MACE distribution (down), demonstrated an overall trend where higher predicted probabilities correlate with higher observed MACE rates. Bin count was 15.

## 4. Discussion

This study presents a ML framework that achieved state-of-the-art performance in predicting MACE through the integration of structured multimodal data. The ALL model, incorporating a broad spectrum of parameters, achieved an AUC-ROC of 0.883 ± 0.012, reflecting a significant improvement over established baselines: 30.8% over CAC, 32.2% over PREVENT, and 28.9% over PCE, despite the low MACE prevalence (2.93%). Although it exhibited a lower sensitivity (0.31 ± 0.04) compared to simpler baseline models (0.57-0.60), this trade-off is anticipated due to the low event rate. Importantly, this reduction in sensitivity was accompanied by substantial improvements across other predictive metrics, achieving a favorable balance between precision, specificity, and overall discriminative performance. These findings, offer a reproducible and scalable benchmark for future multimodal research.

While the dataset itself was not the focal point of novelty, its size and diversity add robustness and general relevance to the modeling conclusions.

Our results confirm that single-modality baselines, while clinically endorsed^7,24^ and widely used for risk stratification, provide moderate discriminative power. We demonstrated that model performance increases with the integration of additional clinical, imaging, and laboratory features, supporting the value of multi-source feature accumulation.

CAC, which has been established as an imaging biomarker for cardiovascular risk^25,26^, yielded an AUC-ROC of 0.675 in our cohort. However, our results show that CAC and IMAGE-derived features, when used in isolation, are not sufficient for accurate long-term risk stratification. SHAP analysis reaffirmed that biochemical and metabolic markers, specifically creatinine, HbA1c, and BMI, had the highest overall impact on predictions. These markers, which reflect renal and metabolic health, are also incorporated in the PREVENT score, whose recent updates emphasize the growing recognition of these pathways in MACE risk. Lesion burden (e.g., CAC volume) also ranked among the top contributors, but traditional risk scores were less influential than expected. These patterns suggest that structured clinical and biochemical data hold significant predictive signal that traditional scoring systems may not fully capture.

Our XGBoost model outperformed recent ML studies despite their favorable conditions. Studies by Ghorashi et al.^27^ and Tesche et al.^28^ reported strong AUC-ROC (0.82– 0.96), but both relied on smaller cohorts (<500 patients), shorter follow-up, and higher event prevalence, limiting generalizability. Among larger studies, Pezel et al.^29^ achieved 0.86 AUC-ROC in ∼2,000 higher risk patients with shorter follow-up using two imaging modalities (CT and MRI).

By contrast, our work considerably advances prior research by integrating comprehensive multimodal clinical scores, imaging metrics, biomarkers, and EKG parameters within a large and diverse patient cohort, achieving strong long-term calibration and overall discriminative performance. A notable strength of our work is the extensive demographic and geographic diversity of our cohort, derived from multiple Emory-affiliated hospitals and healthcare facilities spanning several counties throughout Georgia, USA, encompassing significant racial representation. This demographic breadth, combined with diverse imaging modalities, laboratory equipment, and clinical procedures, ensures robust, clinically actionable results with enhanced generalizability and real-world applicability.

Despite these strengths, certain limitations exist. This was a retrospective clinical cohort study, and although the data were sourced from a geographically diverse healthcare system, external validation across national and international multi-center cohorts is needed. Second, while we leveraged imaging-derived metrics, we did not incorporate volumetric CT image data. This limits the model’s ability to learn spatial or morphological features inherent in the imaging itself. Third, although imputation was not performed, certain variables exhibited missingness, which may introduce bias in both feature importance and model calibration if missingness patterns were informative. Future work should systematically address missing data handling and assess whether missingness itself carries prognostic information. Additionally, future work should explore hybrid architectures that combine structured inputs, like the ones used in this study, with volumetric imaging data to enhance predictive performance.

## 5. Conclusion

We present a powerful, interpretable, and statistically rigorous ML model that establishes a robust benchmark for 10-year MACE prediction. Our ALL feature model sets a new baseline for cardiac risk assessment and provides a reproducible foundation for advancing multimodal fusion strategies in cardiovascular AI. Future studies can build on this work by integrating volumetric imaging and external datasets, ultimately moving toward real-world clinical deployment and closing the gap between classical risk prediction tools and future deep multimodal frameworks.

## Data Availability

All data produced in the present study are available upon reasonable request to the authors.

## References

1. Lindstrom, M. et al. Global Burden of Cardiovascular Diseases and Risks Collaboration, 1990-2021. JACC 80, 2372–2425 (2022).

2. 2. Heart Disease and Stroke Statistics—2021 Update | Circulation. https://www.ahajournals.org/doi/10.1161/CIR.0000000000000950.

3. 3. Martin, S. S. et al. 2025 Heart Disease and Stroke Statistics: A Report of US and Global Data From the American Heart Association. Circulation 151, e41–e660 (2025).

4. 4. Virani, S. S. et al. 2023 AHA/ACC/ACCP/ASPC/NLA/PCNA Guideline for the Management of Patients With Chronic Coronary Disease: A Report of the American Heart Association/American College of Cardiology Joint Committee on Clinical Practice Guidelines. Circulation 148, e9–e119 (2023).

5. Medina-Inojosa, J. R. et al. Performance of the ACC/AHA Pooled Cohort Cardiovascular Risk Equations in Clinical Practice. J. Am. Coll. Cardiol. 82, 1499–1508 (2023).

6. Muntner, P. et al. Validation of the Atherosclerotic Cardiovascular Disease Pooled Cohort Risk Equations. JAMA 311, 1406–1415 (2014).

7. Goff, D. C. et al. 2013 ACC/AHA Guideline on the Assessment of Cardiovascular Risk. J. Am. Coll. Cardiol. 63, 2935–2959 (2014).

8. Karmali, K. N., Goff, D. C., Ning, H. & Lloyd, -Jones Donald M. A Systematic Examination of the 2013 ACC/AHA Pooled Cohort Risk Assessment Tool for Atherosclerotic Cardiovascular Disease. JACC 64, 959–968 (2014).

9. Khan, S. S. et al. Development and Validation of the American Heart Association’s PREVENT Equations. Circulation 149, 430–449 (2024).

10. Khan, S. S. et al. Novel Prediction Equations for Absolute Risk Assessment of Total Cardiovascular Disease Incorporating Cardiovascular-Kidney-Metabolic Health: A Scientific Statement From the American Heart Association. Circulation 148, 1982–2004 (2023).

11. Agatston, A. S. et al. Quantification of coronary artery calcium using ultrafast computed tomography. J. Am. Coll. Cardiol. 15, 827–832 (1990).

12. Blaha, M. J., Blankstein, R. & Nasir, K. Coronary Artery Calcium Scores of Zero and Establishing the Concept of Negative Risk Factors∗. JACC 74, 12–14 (2019).

13. Blaha, M. J. et al. Role of Coronary Artery Calcium Score of Zero and Other Negative Risk Markers for Cardiovascular Disease: The Multi-Ethnic Study of Atherosclerosis (MESA). Circulation 133, 849–858 (2016).

14. Detrano, R. et al. Coronary calcium as a predictor of coronary events in four racial or ethnic groups. N. Engl. J. Med. 358, 1336–1345 (2008).

15. McClelland, R. L., Chung, H., Detrano, R., Post, W. & Kronmal, R. A. Distribution of coronary artery calcium by race, gender, and age: results from the Multi-Ethnic Study of Atherosclerosis (MESA). Circulation 113, 30–37 (2006).

16. Motwani, M. et al. Machine learning for prediction of all-cause mortality in patients with suspected coronary artery disease: a 5-year multicentre prospective registry analysis. Eur. Heart J. ehw188 (2016) doi:10.1093/eurheartj/ehw188.

17. Al’Aref, S. J. et al. Machine learning of clinical variables and coronary artery calcium scoring for the prediction of obstructive coronary artery disease on coronary computed tomography angiography: analysis from the CONFIRM registry. Eur. Heart J. 41, 359–367 (2020).

18. Nakanishi, R. et al. Machine Learning Adds to Clinical and CAC Assessments in Predicting 10-Year CHD and CVD Deaths. JACC Cardiovasc. Imaging 14, 615–625 (2021).

19. Nakanishi, R. et al. The relationship between coronary artery calcium score and the long-term mortality among patients with minimal or absent coronary artery risk factors. Int. J. Cardiol. 185, 275–281 (2015).

20. Chen, T. & Guestrin, C. XGBoost: A Scalable Tree Boosting System. in Proceedings of the 22nd ACM SIGKDD International Conference on Knowledge Discovery and Data Mining 785–794 (ACM, San Francisco California USA, 2016). doi:10.1145/2939672.2939785.

21. Winkel, D. J. et al. Deep learning for vessel-specific coronary artery calcium scoring: validation on a multi-centre dataset. Eur. Heart J. Cardiovasc. Imaging 23, 846–854 (2022).

22. Martin, S. S. et al. Evaluation of a Deep Learning–Based Automated CT Coronary Artery Calcium Scoring Algorithm. JACC Cardiovasc. Imaging 13, 524–526 (2020).

23. Lundberg, S. M. & Lee, S.-I. A Unified Approach to Interpreting Model Predictions. in Advances in Neural Information Processing Systems vol. 30 (Curran Associates, Inc., 2017).

24. Arnett, D. K. et al. 2019 ACC/AHA Guideline on the Primary Prevention of Cardiovascular Disease: A Report of the American College of Cardiology/American Heart Association Task Force on Clinical Practice Guidelines. Circulation 140, (2019).

25. Budoff, M. J. et al. Prognostic Value of Coronary Artery Calcium in the PROMISE Study (Prospective Multicenter Imaging Study for Evaluation of Chest Pain). Circulation 136, 1993–2005 (2017).

26. Lee, J. H. et al. The Predictive Value of Coronary Artery Calcium Scoring for Major Adverse Cardiac Events According to Renal Function (from the Coronary Computed Tomography Angiography Evaluation for Clinical Outcomes: An International Multicenter [CONFIRM] Registry). Am. J. Cardiol. 123, 1435–1442 (2019).

27. Ghorashi, S. M. et al. Comparison of conventional scoring systems to machine learning models for the prediction of major adverse cardiovascular events in patients undergoing coronary computed tomography angiography. Front. Cardiovasc. Med. 9, (2022).

28. Tesche, C. et al. Improved long-term prognostic value of coronary CT angiography-derived plaque measures and clinical parameters on adverse cardiac outcome using machine learning. Eur. Radiol. 31, 486–493 (2021).

29. Pezel, T. et al. A Machine Learning Model Using Cardiac CT and MRI Data Predicts Cardiovascular Events in Obstructive Coronary Artery Disease. Radiology 314, e233030 (2025).

